# Sex differences in the diagnosis of advanced cancer and subsequent outcome in people with chronic kidney disease: an analysis of a national population cohort

**DOI:** 10.1101/2023.08.22.23294412

**Authors:** R Shemilt, MK Sullivan, P Hanlon, B Jani, N De La Mata, B Rosales, BMP Elyan, M Wyld, JA Hedley, R Cutting, DA McAllister, AC Webster, PB Mark, JS Lees

## Abstract

**Background:** In the general population, advanced cancer stage at presentation is associated with poorer health outcomes. People with chronic kidney disease (CKD) have increased incidence and mortality from most cancer types. We sought to determine whether people with CKD were more likely to present with advanced stage cancer, whether this was associated with survival, and whether these associations varied by sex.

**Methods:** Data were from Secure Anonymised Information Linkage Databank (SAIL), a Welsh primary care database with linkage to cancer and death registries. We included patients with a de-novo cancer diagnosis (2011-2017), and at least two kidney function tests in the two years prior to diagnosis. Estimated glomerular filtration rate based on serum creatinine (eGFRcr) was calculated using the CKD-EPI 2009 equation (mL/min/1.73m^2^). Logistic regression models determined odds of presenting with advanced cancer (stage 3 or 4 at diagnosis) by different values of eGFRcr at baseline. Cox proportional hazards models tested associations between eGFRcr at baseline and all-cause mortality risk (reference eGFR 75 to <90).

**Findings:** There were 66,128 patients: 30,857 (46.7%) were female, mean age was 69.1 (standard deviation [SD] 13.8) years in females and 70.6 (SD 11.1) years in males; median eGFRcr at baseline was 78 (interquartile range [IQR] 63 – 90) mL/min/1.73m^2^ in both females and males. Over a median follow-up time of 3.1 (IQR 0.5 – 5.7) years in females and 2.9 (IQR 0.5-5.5) years in males, there were 17,303 deaths in females and 20,855 in males. An eGFRcr <30 was associated with higher odds of presenting with advanced cancer in males (OR 1.33 95% CI 1.09-1.62), but not in females (OR 1.17 95% CI 0.92-1.50); positive associations were primarily driven by prostate and breast cancers. With lower eGFRcr, hazards of cancer death increased in both sexes, but lower eGFRcr was associated with greater hazards of cancer death in females (eGFRcr <30: HR 1.71, 95% CI 1.56-1.88, p<0.001; male versus female comparison HR 0.88, 95% CI 0.78-0.90; p=0.037).

**Interpretation:** CKD was not associated with substantially higher odds of presenting with advanced cancer across most cancer sites (except prostate and breast), but was associated with reduced survival. Despite an initial survival advantage compared to males, females with CKD had disproportionately higher hazards of death. Though potential explanations for reduced survival after a cancer diagnosis are manifold, scrutiny of access to, efficacy, and safety of cancer treatments in people with CKD – particularly females with CKD – are warranted.

**Funding:** Chief Scientist Office (Scotland) Postdoctoral Lectureship (PCL/20/10) and University of Sydney/University of Glasgow Office of Global Engagement Collaboration Partnership (9241562498).

## Introduction

Chronic kidney disease (CKD) poses a significant healthcare burden globally: CKD affects approximately 11-13% of the population^1^, and is becoming more common, driven by ageing and multi-morbidity^2^. Due to shared risk factors, CKD is more common among people with other comorbid diseases, particularly cardiovascular disease and cancer. Depending on the cancer site, CKD may be present in up to 50% of people diagnosed with cancer^3^. Furthermore, there are important sex differences in cancer outcomes: in the general population, females have better survival from cancer than males; however, females with CKD and kidney failure have worse relative survival, more excess deaths, and more years of life lost to cancer^4,5^. The loss of female survival advantage in CKD is not well understood.

With increasing severity of CKD (reduced estimated glomerular filtration rate [eGFR] and albuminuria), the risk of cancer death rises^6–8;^ however, the mechanisms are uncertain. In the general population, diagnosis of cancer at a more advanced cancer stage results in poorer cancer outcomes^9,10^: Curative treatment options are more limited with advanced staging^9,11^. The presence of CKD may further restrict access to, safety and/or efficacy of cancer treatments, including surgery with curative intent and systemic anti-cancer therapies^12,13^. However, CKD may also influence presenting cancer stage: there may be differences in cancer biology, in the timing and nature of healthcare interactions and/or in the investigation and/or management of non-specific symptoms seen commonly in both CKD and cancer (e.g. anaemia and weight loss). It is conceivable that presentation with more advanced cancer stage explains reduced survival after a cancer diagnosis among people with CKD; however, this has not previously been investigated.

In the general population, sex differences in cancer incidence and outcome are well-documented^14^. Predominant cancer sites and associated prognosis vary considerably in people of male and female sex, with a significant impact on overall sex differences in cancer outcomes^15^. In some cases this is due to obvious anatomical, hormonal or epidemiological differences. Less is known about other factors which may influence differences in cancer outcome between sexes - such as timing of presentation and variation in treatment strategy - and whether such differences explain the loss of female survival advantage among people with CKD.

Using data from a large primary care cohort, we sought to identify whether patients with CKD were more likely to present with advanced cancer, whether more invasive stage at presentation was associated with reduced survival in CKD, and whether these factors varied by sex.

## Methods

### Data sources and population

Data were from Secure Anonymised Information Linkage Databank (SAIL): a Welsh primary care database with linkage to cancer (Wales Cancer Intelligence and Surveillance Unit; WCISU) and death (Office for National Statistics; ONS) registries. Participants were included if they had: i) a de-novo diagnosis of malignant cancer between 1^st^ January 2011 to 31^st^ December 2017 (by International Classification of Disease (ICD)-10 code C00-C75 (excluding C44 – as reporting of non-melanoma skin cancers are not mandated in cancer registries); and ii) if they had kidney function tested at least twice, at least three months apart, and within two years prior to the cancer diagnosis. Participants were excluded if they were receiving maintenance kidney replacement therapy (KRT) at the time of cancer diagnosis.

As a measure of kidney function, estimated glomerular filtration rate was calculated from serum creatinine without including the race coefficient (eGFRcr; CKD-EPI 2009 equation^16^), as this is the method currently recommended for use in UK populations^17^. In keeping with many populations with CKD in primary care, albuminuria was not consistently available for CKD staging^18^.

Participant demographics were extracted from the primary care record. Age was calculated in years between date of birth and date of first cancer diagnosis in the period of interest. Sex was recorded in the clinical record as “male” or “female”. Smoking status was coded as “never smoker”, “ex-smoker”, or “current smoker”. Comorbidites were defined according to a previously published list of 40 long-term conditions^19^, defined using Read Codes from primary care records as previously described^20,21^. Comorbidity count was calculated as the sum of the number of long-term conditions, excluding CKD and cancer. Deprivation status was expressed using the Welsh Index of Multiple Deprivation (WIMD) 2011^22^, which considers 8 weighted indices (income, employment, health, education, geographical access to services, housing, physical environment and community safety) according to home postcode to provide a ranked WIMD score. WIMD was expressed in deciles from 1 (most deprived) to 10 (least deprived).

For site-specific analyses, cancer site was determined from ICD-10 codes as the first cancer in the follow-up period. A full list of groupings by cancer site is available in Table 1. For site-specific analyses, we excluded cancers where there were fewer than 500 diagnoses in the total population for reasons of patient confidentiality, and to avoid invalid statistical inference. This excluded people with a first cancer of the male genital organs, bone, thyroid, adrenal, endocrine and brain/central nervous system cancers from further analysis.

**Table 1.** Baseline data.

|  | All | Female | Male | P value |
| --- | --- | --- | --- | --- |
| **N (%)** | 66,128 (100) | 30,857 (46.7) | 35,271 (53.3) |  |
| **Age (years): mean (SD)** | 69.9 (12.5) | 69.1 (13.8) | 70.6 (11.1) | <0.001 |
| **eGFR (mL/min/1.73m<sup>2</sup>): median [IQR]** | 78.5 [63.0, 89.8] | 78.3 [62.9, 90.0] | 78.5 [63.2, 89.7] | 0.543 |
| **eGFR category (mL/min/1.73m<sup>2</sup>): n(%)** |  |  |  | <0.001 |
| **eGFR >120** | 21,026 (31.8) | 9,630 (31.2) | 11,396 (32.3) |  |
| **eGFR 105 - <120** | 283 (0.4) | 152 (0.5) | 131 (0.4) |  |
| **eGFR 90 - <105** | 2,290 (3.5) | 1,234 (4.0) | 1,056 (3.0) |  |
| **eGFR 75 - <90** | 13,680 (20.7) | 6,327 (20.5) | 7,353 (20.8) |  |
| **eGFR 60 - <75** | 14,641 (22.1) | 6,849 (22.2) | 7,792 (22.1) |  |
| **eGFR 45 - <60** | 8,069 (12.2) | 3,795 (12.3) | 4,274 (12.1) |  |
| **eGFR 30 - <45** | 4,428 (6.7) | 2,133 (6.9) | 2,295 (6.5) |  |
| **eGFR <30** | 1,711 (2.6) | 737 (2.4) | 974 (2.8) |  |
| **Smoking status: n (%)** |  |  |  | <0.001 |
| **Current smoker** | 13,729 (20.8) | 6,426 (20.8) | 7,303 (20.7) |  |
| **Ex smoker** | 19,847 (30.0) | 7,244 (23.5) | 12,603 (35.7) |  |
| **Non smoker** | 21,348 (32.3) | 11,438 (37.1) | 9,910 (28.1) |  |
| **Missing** | 11,204 (16.9) | 5,749 (18.6) | 5,455 (15.5) |  |
| **Comorbidity count: median [IQR]** | 2 [1, 4] | 2 [1, 4] | 2 [1, 4] | <0.001 |
| **WIMD decile: median [IQR]** | 5 [3, 8] | 5 [3, 8] | 6 [3, 8] | <0.001 |
| **Cancer stage: n (%)** |  |  |  | <0.001 |
| **1 (Least advanced)** | 13,277 (20.1) | 8,162 (26.5) | 5,115 (14.5) |  |
| **2** | 13,118 (19.8) | 5,663 (18.4) | 7,455 (21.1) |  |
| **3** | 10,900 (16.5) | 4,703 (15.2) | 6,197 (17.6) |  |
| **4 (Most advanced)** | 15,371 (23.2) | 6,089 (19.7) | 9,282 (26.3) |  |
| **Unknown** | 13,462 (20.4) | 6,240 (20.2) | 7,222 (20.5) |  |

### Outcomes

We were interested in the following outcomes:

1. Presentation with advanced cancer; i.e., stage 3 or 4 cancer by Tumour Node Metastases (TNM), numeric grading systems, or – for female genital organ cancers – International Federation of Gynecology and Obstetrics (FIGO).
2. Death (from any cause) after cancer diagnosis during the follow-up period.

### Statistical analysis

Data summaries are stratified by sex and expressed as mean (standard deviation: SD), median (interquartile range: IQR) and count (%), and compared using t-test, Kruskal-Wallis and chi-squared test as appropriate.

To determine odds of presenting with advanced cancer by eGFR (overall, and by cancer site), we applied logistic regression models, adjusted for age, deprivation status, smoking status, comorbidity count plus cancer site (for overall, but not site-specific models). Where staging information was unavailable for other solid organ cancers, presenting cancer stage was allocated as “unknown” (where stage was recorded as “GX” or where no staging information was recorded) according to data collected within the cancer registry.

To determine hazards of death (from any cause; overall, and by cancer site) after cancer diagnosis by eGFR, we constructed Cox proportional hazards models adjusted for age, deprivation status, smoking status, comorbidity count, cancer site (for overall, but not site-specific models) and cancer stage at presentation. Follow-up was from cancer diagnosis until the sooner of date of death or 1^st^ October 2020.

In order to describe the potential associations at different stages of CKD, eGFR was categorised in 15 mL/min/1.73m^2^ decrement (using the two most recent eGFR measurements, taken at least 3 months apart and within 2 years prior to cancer diagnosis) as follows: eGFR >120, eGFR >105 – 120, eGFR >90 – 105, eGFR >75 – 90 (reference), eGFR >60 – 75, eGFR >45 – 60, eGFR >30 – 45, eGFR <30. Where there smaller numbers at eGFR extremes (e.g. testing associations), the top two and bottom two categories were collapsed. eGFR categories for site-specific cancers were therefore as follows: eGFR >105, eGFR >90 – 105, eGFR >75 – 90 (reference), eGFR >60 – 75, eGFR >45 – 60, eGFR <= 45.

Evidence of a statistical interaction was sought between sex and eGFR category in both logistic regression and Cox proportional hazards models. Results are presented: i) stratified by sex and ii) indicating where significant interactions exist between sex and eGFR. To account for a substantial proportion of included patients who had missing cancer stage, sensitivity analyses (assuming highest or lowest possible stage) were conducted.

Analyses were conducted using *tidyverse, nephro, broom, tableone* and *survival* packages for R statistical software (version 4.1.3).

## Results

Of 141,784 patients with a new diagnosis of cancer, there were 66,128 with two available kidney function measures for CKD staging who were included in the analyses. People with cancer who were excluded due to insufficient kidney function tests to meet eligibility criteria were younger, with similar median eGFRcr (based on single measure alone), comorbidity count and deprivation status, but slightly lower proportion of current smokers (Supplementary Table S1). There was a slightly higher preponderance of females in this group; however, sex was not recorded for 36.1%.

In our included cohort, 46.7% were female, with mean age in females 69.1 (SD 13.8) years and in males 70.6 (SD 11.1) years (Table 1). There were 14,208 individuals (21.5% overall; 21.6% in females and 21.4% in males) with moderate-advanced CKD (eGFR <60mL/min/1.73m^2^) at baseline. Over median follow-up time 3.1 (IQR 0.5 to 5.7) years in females and 2.9 (IQR 0.5 to 5.5) years in males, there were 17,303 deaths in females and 20,855 deaths in males. Median survival times for site-specific cancers in males and females were shortest for abdominal and respiratory cancers, and longest for melanoma in both males and females (Table 2).

**Table 2.**
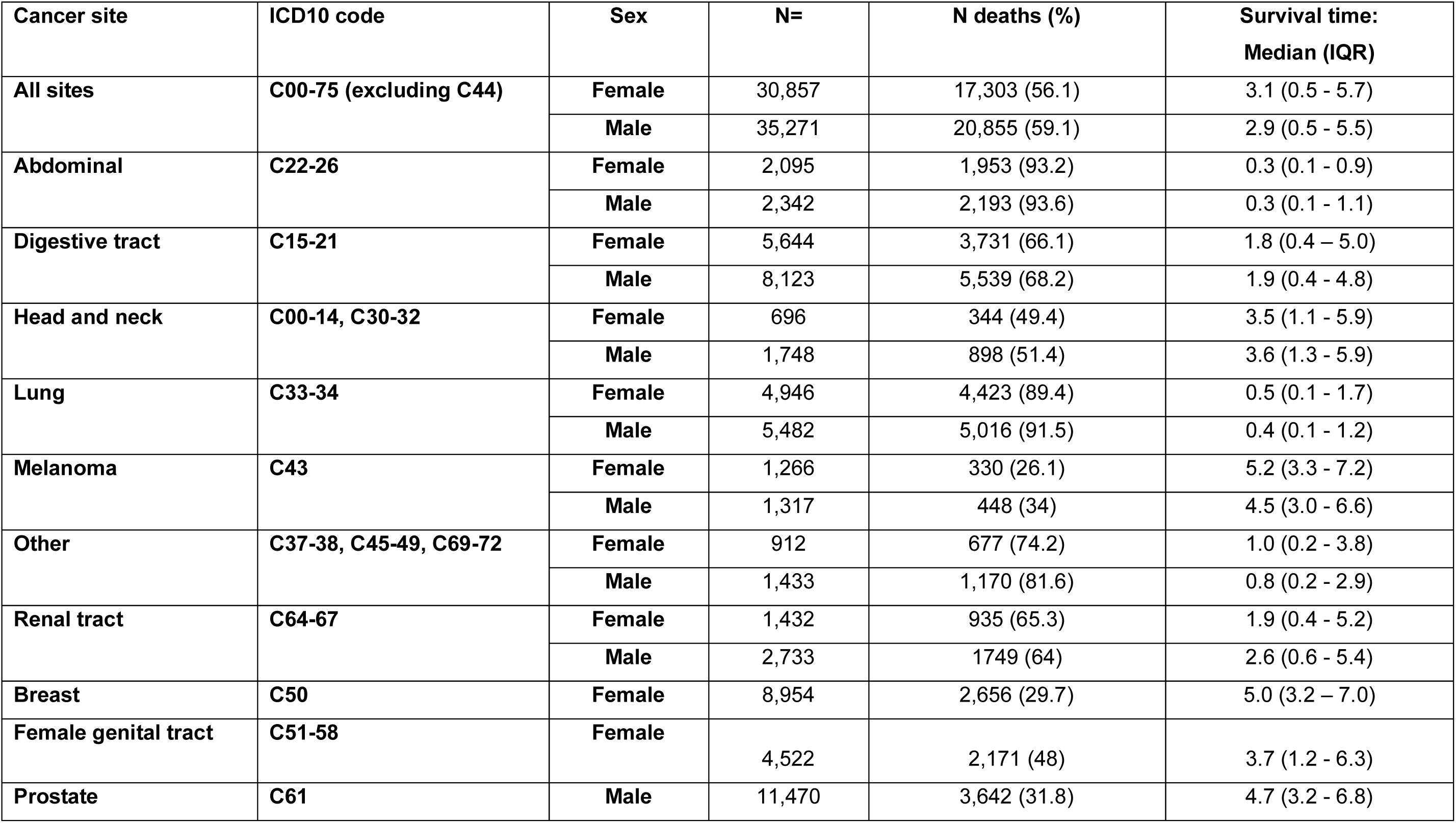
Overall and site-specific cancer diagnoses, deaths and survival times in females and males.

| Cancer site | ICD10 code | Sex | N= | N deaths (%) | Survival time:<br>Median (IQR) |
| --- | --- | --- | --- | --- | --- |
| All sites | C00-75 (excluding C44) | Female | 30,857 | 17,303 (56.1) | 3.1 (0.5 - 5.7) |
|  |  | Male | 35,271 | 20,855 (59.1) | 2.9 (0.5 - 5.5) |
| Abdominal | C22-26 | Female | 2,095 | 1,953 (93.2) | 0.3 (0.1 - 0.9) |
|  |  | Male | 2,342 | 2,193 (93.6) | 0.3 (0.1 - 1.1) |
| Digestive tract | C15-21 | Female | 5,644 | 3,731 (66.1) | 1.8 (0.4 – 5.0) |
|  |  | Male | 8,123 | 5,539 (68.2) | 1.9 (0.4 - 4.8) |
| Head and neck | C00-14, C30-32 | Female | 696 | 344 (49.4) | 3.5 (1.1 - 5.9) |
|  |  | Male | 1,748 | 898 (51.4) | 3.6 (1.3 - 5.9) |
| Lung | C33-34 | Female | 4,946 | 4,423 (89.4) | 0.5 (0.1 - 1.7) |
|  |  | Male | 5,482 | 5,016 (91.5) | 0.4 (0.1 - 1.2) |
| Melanoma | C43 | Female | 1,266 | 330 (26.1) | 5.2 (3.3 - 7.2) |
|  |  | Male | 1,317 | 448 (34) | 4.5 (3.0 - 6.6) |
| Other | C37-38, C45-49, C69-72 | Female | 912 | 677 (74.2) | 1.0 (0.2 - 3.8) |
|  |  | Male | 1,433 | 1,170 (81.6) | 0.8 (0.2 - 2.9) |
| Renal tract | C64-67 | Female | 1,432 | 935 (65.3) | 1.9 (0.4 - 5.2) |
|  |  | Male | 2,733 | 1749 (64) | 2.6 (0.6 - 5.4) |
| Breast | C50 | Female | 8,954 | 2,656 (29.7) | 5.0 (3.2 – 7.0) |
| Female genital tract | C51-58 | Female | 4,522 | 2,171 (48) | 3.7 (1.2 - 6.3) |
| Prostate | C61 | Male | 11,470 | 3,642 (31.8) | 4.7 (3.2 - 6.8) |

### Comparison of males versus females without accounting for kidney function

Adjusted for age alone, males were more likely than females to present with advanced cancer (odds ratio: OR 1.51, 95% CI 1.47 – 1.57; p<0.001). The association was attenuated but preserved in fully adjusted models (OR 1.15, 95% CI 1.09 – 1.22; p<0.001). Adjusted for age alone, males had similar risk of cancer death compared to females (hazard ratio: HR 1.01, 95% CI 0.99-1.03; p=0.48); however, in fully adjusted models (including adjustments for cancer site and cancer stage at presentation but not accounting for eGFR category), males were at increased risk of all-cause mortality after a cancer diagnosis than females (HR 1.11 95% CI 1.08-1.13; p<0.001). Without accounting for kidney function, these findings show that females had a survival advantage compared to males.

### Risk of presenting with advanced cancer in CKD

In both males and females, there were very small increased odds of presenting with advanced cancer when all sites were included in people with extremes of eGFR (Supplementary Table S2 and Figure 1A). This was most pronounced at very low eGFR (<30: reflecting CKD stage 4-5) and very high eGFR (>105-120; >120: not currently considered to reflect presence of CKD) in males. In females, OR for presenting with advanced cancer with low eGFR (<30; 30 - <45) and high eGFR (>105-120; >120) crossed the null. There was a statistical interaction between eGFR and sex at high levels of eGFR: males were at higher odds of presenting with advanced cancer (105 - <120: OR 1.35, 95% CI 1.03-1.78; p=0.03; >=120: OR 3.27, 95% CI 1.41-7.97; p=0.01; Figure 1A).

**Figure 1.**
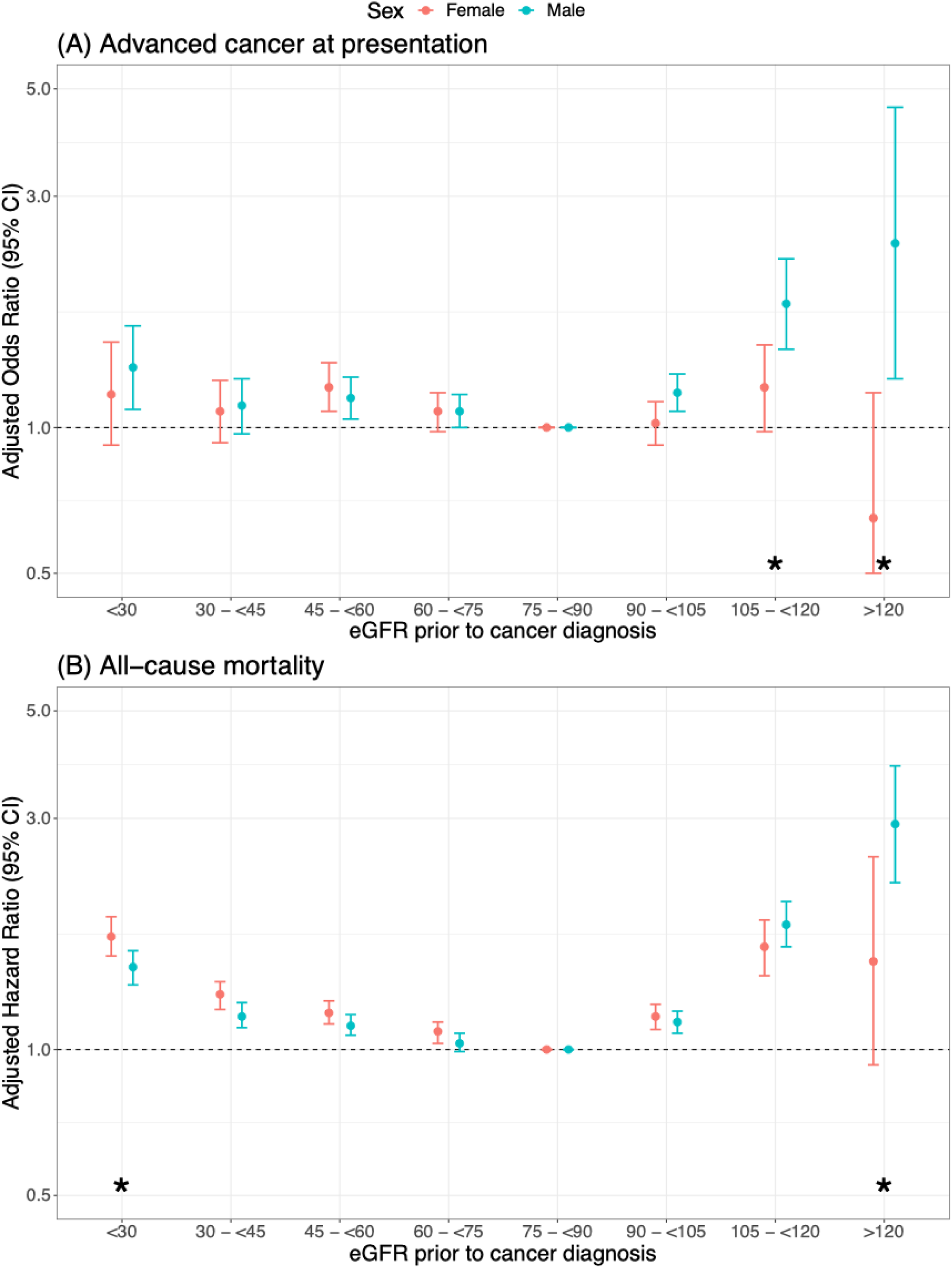
**(A)** Plot displaying OR (95% CI) of presentation with advanced cancer. Models are adjusted for age, sex, smoking status, deprivation status, number of comorbidities. **(B)** Plot displaying HR (95% CI) of death after cancer diagnosis. Models are adjusted for age, sex, smoking status, deprivation status, number of comorbidities and presenting cancer stage. Results are stratified by sex. * indicates presence of a significant interaction between sex and eGFR category. Reference eGFR category: 75 - <90 mL/min/1.73m^2^.

The likelihood of presenting with advanced site-specific cancer also differed by sex and eGFR (Supplementary Table S3 and Figure 2). In females, people with very low eGFR (<45) or very high eGFR (>=105) were more likely to present with advanced breast cancer. A similar finding was seen in males with prostate cancer. There was not a significant association between lower eGFR and likelihood of presenting with advanced cancer across any other solid organ cancer site. Very high eGFR (>=105) was associated with increased likelihood of presenting with advanced cancer in digestive tract cancers (in males) and lung cancers (in females).

**Figure 2.**
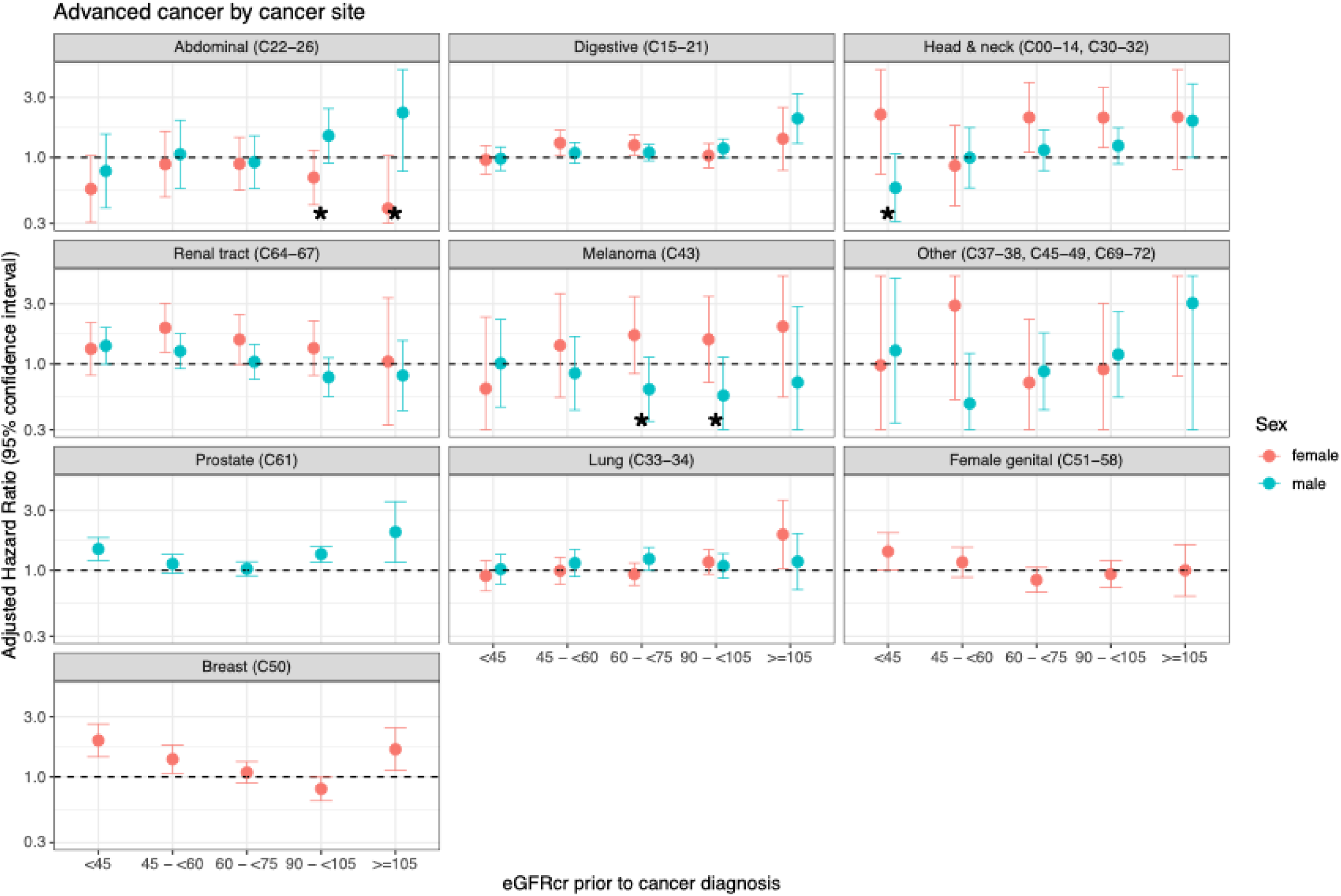
Plot displaying odds ratio (OR; 95% confidence intervals) of presentation with advanced (stage 3 or 4) site-specific cancer. Models are adjusted for age, sex, smoking status, deprivation status and number of comorbidities. Results are stratified by sex and cancer site. * indicates presence of a significant interaction between sex and eGFR category.

### All-cause mortality after cancer diagnosis in CKD

In both males and females, adjusted hazards of death after a cancer diagnosis were higher with eGFR both lower (<75) and higher (≥90) than the reference category; the pattern was more pronounced at extremes of eGFR (Supplementary Tables S4 and Figure 1B). There was a statistical interaction between eGFR and sex at eGFR <30 (males had lower hazards of cancer death than females: HR 0.88, 95% CI 0.78-0.99; p=0.04) and at eGFR >120 (males had higher hazards of cancer death than females: HR 1.80, 95% CI 1.02-3.16; p=0.04; Supplementary Table S4 and Figure 1B). On sensitivity analyses, assuming the maximum and minimum possible cancer stage for those with missing information, findings were similar (data available on request).

In site-specific cancers, low eGFR (<45) was associated with higher hazards of death in people diagnosed with abdominal organ cancers (females more than males), digestive tract cancers (females more than males), with similar higher hazards in males and females for haematological cancers including myeloma, renal tract, lung and non-melanoma skin cancers, prostate (in males only) and breast (females only) cancers (Supplementary Table S5 and Figure 3). High eGFR (>=105) was associated with higher hazards of death in both males and females with abdominal organ, digestive tract, head and neck, melanoma, non-melanoma skin, lung, breast (females only) and prostate (males ony) cancers (Figure 3).

**Figure 3.**
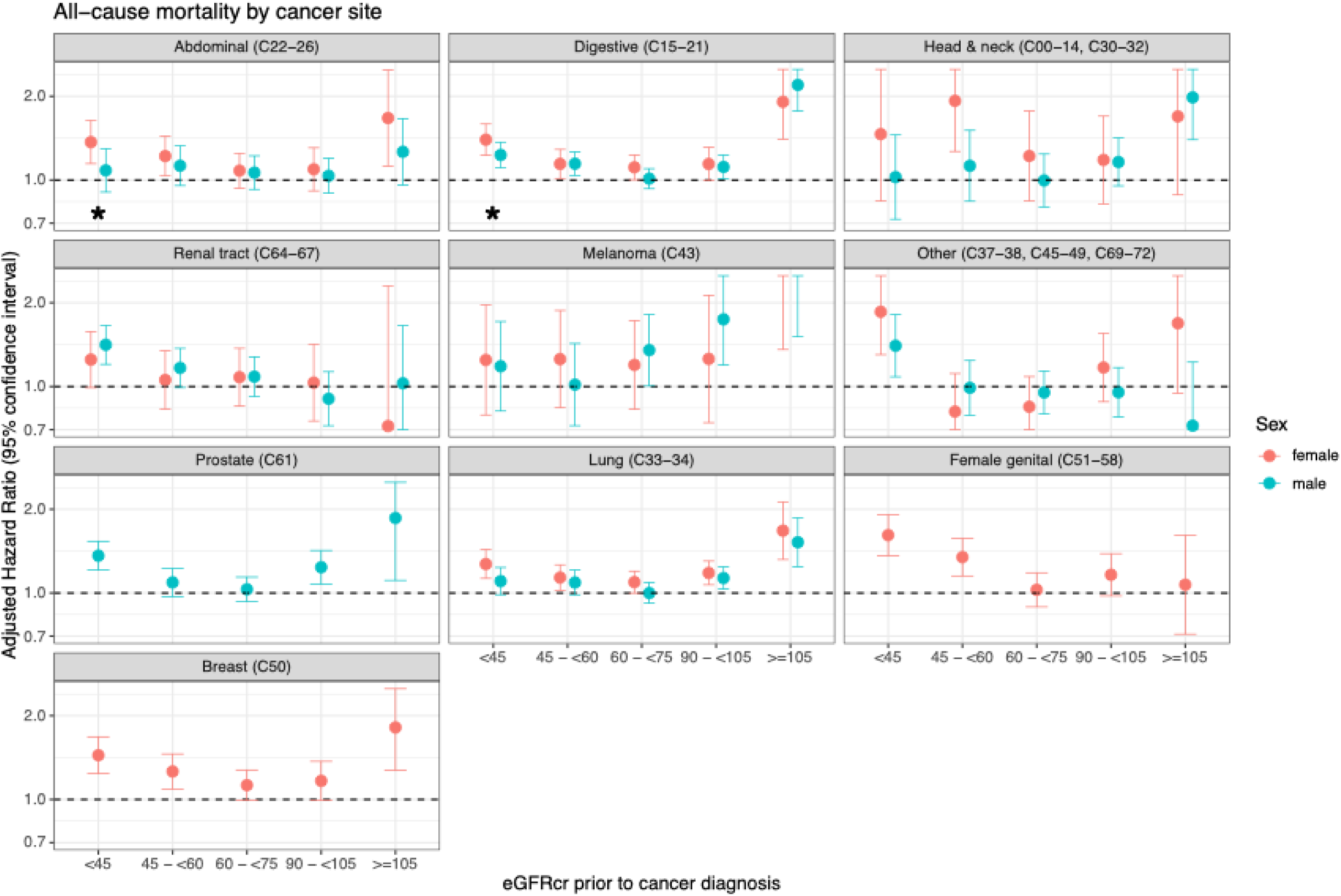
Plot displaying hazards ratios (HR; 95% confidence intervals) of death (any cause) after site-specific cancer diagnosis. Models are adjusted for age, sex, smoking status, deprivation status, number of comorbidities and presenting cancer stage. Results are stratified by sex and cancer site. * indicates presence of a significant interaction between sex and eGFR category.

## Discussion

In this large primary care cohort, people with CKD had higher odds of presenting with advanced cancer of the breast and prostate, but not with advanced cancer at other sites. Despite this, lower (and higher) eGFR was associated with increased hazards of death following cancer diagnosis, suggesting that the excess life lost to cancer in CKD may be linked to differences in post-diagnosis cancer care. The associations between lower eGFR and outcomes varied according to cancer site and sex: females with CKD particularly lost their survival advantage compared to males in cancers of the abdominal organs and digestive tract.

We are not aware of any prior studies that have examined sex differences in presenting cancer stage and how this may be affected by coexisting CKD. However, our study is in keeping with several prior analyses that show higher hazards of death associated with cancer in people with CKD^4,23–26^. Our data additionally suggest that in people with CKD, there are notable sex differences in outcomes post-cancer diagnosis across several cancer sites. One prior study has also identified that CKD is associated with more years of life lost to cancer in females than in males^4^. To our knowledge, ours is the first study to report that these sex differences in people with CKD still exist after accounting for both the cancer site and stage at presentation.

### Cancer survival difference in CKD

The differences in cancer survival between CKD and non-CKD populations with cancer suggest that differences in post-diagnosis care in CKD, including differential effects of cancer treatment, may explain some of the difference in reduced survival in patientds with CKD and cancer. Due to widespread exclusion from trials of systemic anti-cancer therapies (SACT)^27,28^, there is a paucity of evidence of the efficacy and safety of SACT among people with CKD^29^. However, most anti-cancer drugs are administered near the maximum tolerated dose and have a narrow therapeutic index^30^. Dosing considerations (and kidney function) are therefore particularly important.

Cytotoxic agents including platinum-based chemotherapy (such as carboplatin) and alkylating agents (such as ifosfamide) may cause a number of renal complications including acute tubular injury leading to chronic tubulointerstitial fibrosis^31^. Pre-existing CKD makes these complications more likely and their consequences potentially more serious, meaning that these agents are often avoided altogether in this group^32^.

The use of targeted SACT in cancer management is now widespread. These have achieved improved outcomes in the general population through targeting specific immune mediators and avoiding many of the toxic systemic effects of cytotoxic chemotherapies^13^. Many targeted agents primarily undergo hepatic metabolism: no dose adjustment is expected even in advanced CKD^30^ and there is some evidence from case series of SACT being given safely even to patients on KRT^33^. However, improvements in cancer survival seen in the general population have not been matched in people with CKD, and it is unclear: i) to what extent newer SACT are used in people across the disease spectrum of CKD, and ii) whether SACT efficacy and safety profiles are similar in CKD as in the general population^30^. Given that CKD is common in people with cancer, a better understanding of SACT use in patients with CKD (both in trials and in the post-licensing period) is essential to improve the provision of evidence-based cancer care to people with CKD.

### Sex differences in cancer survival

Differences in treatment selection, efficacy and safety profiles may explain reduced relative survival in female compared with male patients with advanced CKD. A recent meta-analysis of sex differences in cancer immunotherapy efficacy showed a significantly greater relative reduction in risk of death in males treated with immunotherapy compared to females^34^. This analysis also highlighted disparities in the current evidence base for cancer therapies in males and females, with males comprising two-thirds of included participants in the 20 randomised controlled trials. Though the inclusion of females in trials has increased since the 1993 reversal of previous Food and Drug Administration (FDA) guidelines banning females of child-bearing potential from participation in clinical research, male participants continue to predominate^35^. There is a growing case that trial evidence should be interpreted and applied to clinical practice after taking patient sex into account. This suggests that review of the efficacy and safey profiles of cancer therapies in females (particularly females with CKD) is particularly urgent.

Beyond differences in treatment, potential reasons for sex differences in cancer outcomes include differences in environmental exposures, gene expression, immunity and hormones^36^.

The effects of hormones on innate and adaptive immune responses are increasingly recognised, with oestradiol thought to enhance both cell-mediated and humoral immune responses^37^. This has been postulated to be one factor contributing to the increased incidence of autoimmune disease in females and cancer in males. This is of particular interest in the context of kidney disease and cancer: advancing kidney disease impairs function of the hypothalamic-pituitary-gonadal axis and results in failure of oestradiol levels to peak normally mid-menstrual cycle^38^. Low oestradiol has been associated with worsening kidney function^38^. While the specific mechanisms remain unclear, oestradiol may have an immunomodulatory effect which could contribute to sex differences in cancer incidence, outcome and response to treatment. Though beyond the scope of this study, further investigation is required to understand if differences in cancer biology underpin poorer cancer outcome in females (compared to males) with CKD.

### Non-linear relationship between eGFRcr and survival

There was a notable ‘J-shaped’ relationship between eGFRcr and hazards of death after cancer diagnosis, with eGFRcr greater than 90mL/min/1.73m^2^ also associated with poorer outcome: a consistent finding in analyses in other populations^26,39,40^. However, eGFR calculated using an alternative marker of kidney function (cystatin C – not routinely tested or available for comparison in this population) shows a more biologically plausible, linear association between eGFR and cancer death^26^, suggesting that the J-shaped relationship with eGFRcr reflects intrinsic flaws in creatinine-based estimation of kidney function. Muscle mass contributes to systemic error in estimation of eGFRcr which is most significant at extremes, particularly in patients with cachexia, sarcopenia and high muscularity^41^. Patients at higher extremes of eGFRcr may actually reflect worse kidney function, where low muscle mass results in over-estimation of eGFRcr, when in fact low body weight or sarcopenia (common in people with more advanced cancer) may place them in a higher risk group for treatment toxicity and poor outcome^42^. In this situation, the association between higher eGFRcr and worse outcome may reflect reverse causality.

### Strengths and limitations

The strengths of this study lie in the capture of nationally-representative data in people diagnosed with cancer, using cancer registry data to confirm cancer diagnoses, and biochemical (as compared with clinical coding) confirmation of CKD. We acknowledge several limitations. First, we have included only patients who had at least two measures of kidney function in advance of cancer diagnosis, disproportionately collecting information on people with reasons to seek regular medical attention. This represents approximately half of all people diagnosed with cancer over the same time period; however, this is a robust method for the diagnosis of CKD^17^. Kidney function is commonly tested in community populations, and especially in older people, those with long-term conditions or in those with symptoms that might be in keeping with cancer. We showed that our selected population was slightly older are more likely to be current smokers than the unselected group, but with similar eGFRcr, number of comorbidities and deprivation status. Our approach has likely captured a substantial proportion of the kidney disease population. Second, CKD may be caused by certain types of cancer (such as renal tract cancers and myeloma), introducing the possibility of reverse causality. However, our findings were also preserved across a variety of cancer sites in which reverse causality is implausible. Third, in keeping with challenges seen in national registries worldwide, cancer stage was unknown for around 20% of participants. Missing stage may have biased the potential association between likelihood of presenting with advanced cancer in either direction. However, findings were similar whether investigating this association in those with complete information and on sensitivity analyses assuming highest or lowest possible stage.

### What next?

Existing data may provide valuable information on the utility of cancer treatments in female and male patients with CKD. Trial data may be limited by lack of representativeness; routinely collected data may be limited by confounding by indication. We propose detailed exploration of trial and linked routinely collected data cancer treatment in female and male patients with CKD. The aims should be to understand – in people with CKD – if, how and why the following differ compared with people without CKD:

- cancer treatment selection: operative management, radiotherapy, systemic anti- cancer therapy, conservative management
- treatment delivery: time-to-treatment, dose, duration
- efficacy: progression-free survival, overall survival
- safety: serious adverse events, hospitalisations
- clinical trial enrolment

## Conclusions

Chronic kidney disease was associated with higher odds of presenting with advanced cancer of the breast and prostate, but not with higher odds of advanced cancer in other sites. CKD is associated with reduced survival in people diagnosed with cancer. Despite an initial survival advantage compared to males, females with advanced CKD had disproportionately higher hazards of death. Lack of evidence and guidance for cancer treatment in people with CKD

may underpin these findings, and augment sex differences. Particularly in cancer types where sex discrepancies exist (abdominal organ and digestive tract cancers), scrutiny of the selection, delivery, efficacy and safety of cancer treatment in people with CKD is warranted.

## Data Availability

Data are publically available on application to Secure Anonymised Information Linkage Wales (SAIL; https://saildatabank.com). The current analysis was conducted under project 1214. Model outputs and analysis code will be made available at time of publication (https://github.com/UoGSCMHDataScience/sail_cancer_ckd_public/).

https://saildatabank.com

## Supplementary data

**Table S1.**
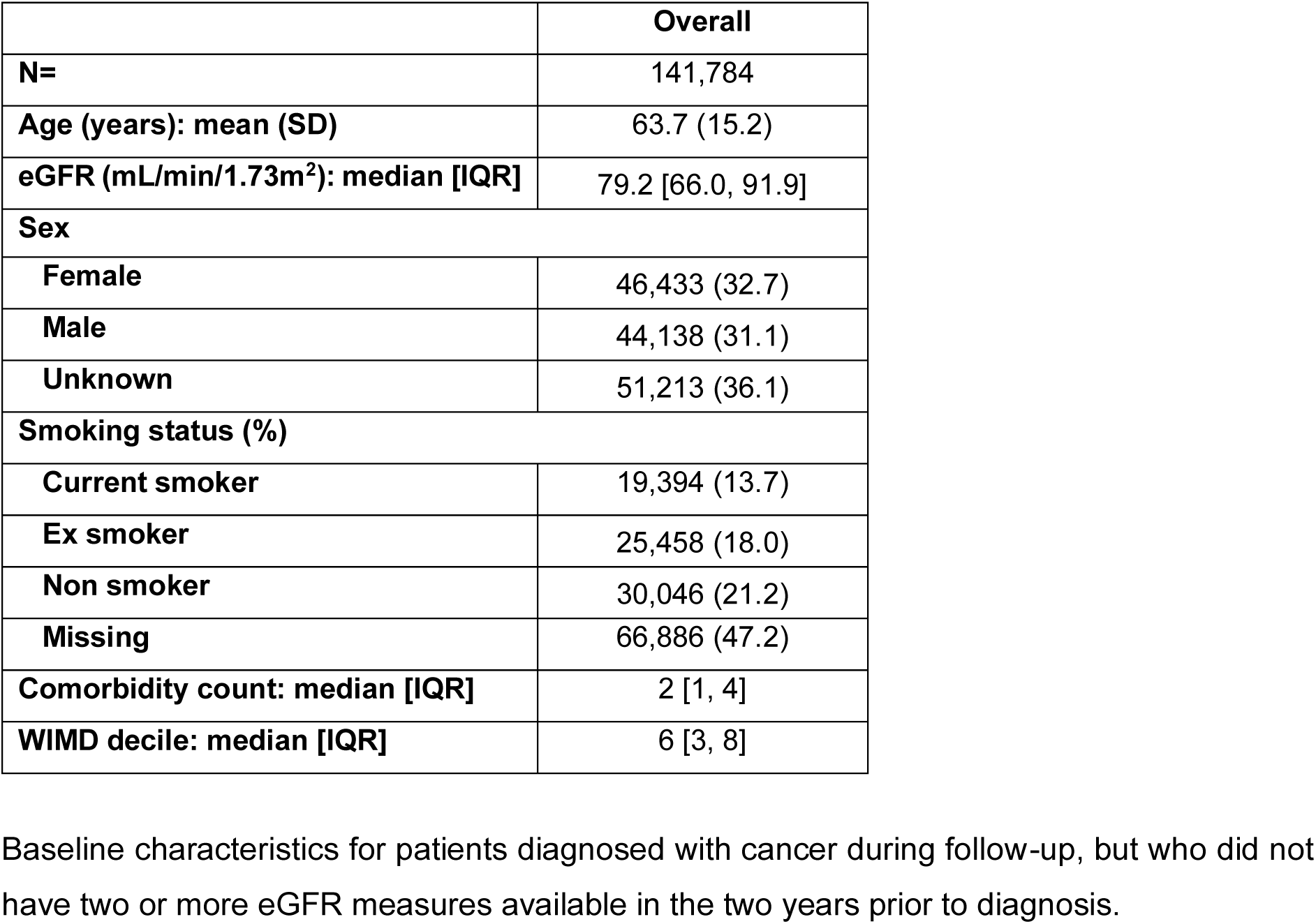
Baseline characteristics for patients diagnosed with cancer during follow-up, but who did not have two or more eGFR measures available in the two years prior to diagnosis.

**Table S2.** Odds of presenting with advanced cancer across all sites by eGFR category.

|  | Female |  | Male |  | Male versus female |  |
| --- | --- | --- | --- | --- | --- | --- |
| eGFR category<br>(mL/min/1.73m<sup>2</sup>) | OR (95% CI) | P value | OR (95% CI) | P value | OR (95% CI) | P value |
| **P for trend** |  | Linear 0.06<br>Non-linear 0.42 |  | Linear 0.089<br>Non-linear <0.001 |  | Linear 0.08<br>Non-linear 0.67 |
| **>120** | 0.65 (0.35-1.18) | 0.15 | 2.40 (1.26-4.58) | 0.008 | 3.27 (1.41-7.97) | 0.01 |
| **105 - <120** | 1.21 (0.98-1.48) | 0.08 | 1.80 (1.45-2.23) | <0.001 | 1.35 (1.03-1.78) | 0.03 |
| **90 - <105** | 1.02 (0.92-1.13) | 0.69 | 1.18 (1.08-1.29) | <0.001 | 1.12 (0.99-1.26) | 0.09 |
| **75 - <90** | Ref | NA | Ref | NA | Ref | NA |
| **60 - <75** | 1.08 (0.98-1.18) | 0.12 | 1.08 (1.00-1.17) | 0.06 | 1.01 (0.89-1.14) | 0.86 |
| **45 - <60** | 1.21 (1.08-1.36) | 0.001 | 1.15 (1.04-1.27) | 0.01 | 0.95 (0.82-1.10) | 0.49 |
| **30 - <45** | 1.08 (0.93-1.25) | 0.31 | 1.11 (0.97-1.26) | 0.13 | 1.02 (0.85-1.24) | 0.81 |
| **<30** | 1.17 (0.92-1.50) | 0.20 | 1.33 (1.09-1.62) | 0.004 | 1.13 (0.83-1.53) | 0.45 |

Logistic regression models adjusted for age, deprivation status, smoking status, comorbidity count and cancer site. eGFR: estimated glomerular filtration rate based on CKD-EPI 2009 equation and using serum creatinine. “Female” and “Male” models are stratified by sex. “Male versus female” model includes an interaction term between eGFR category and sex. Linear: P for linear trend. Non-linear: P for cubic trend. OR: odds ratio. CI: confidence interval.

**Table S3.** Odds of presenting with advanced cancer by cancer site and eGFR category.

|  |  | Female |  | Male |  | Male versus female |  |
| --- | --- | --- | --- | --- | --- | --- | --- |
| Site | eGFR category (mL/min/1.73m<sup>2</sup>) | OR (95% CI) | P value | OR (95% CI) | P value | OR (95% CI) | P value |
| **Abdominal** | **P for trend** |  | Linear 0.94<br>Non-linear 0.03 |  | Linear 0.15<br>Non-linear 0.20 |  | Linear 0.13<br>Non-linear 0.01 |
| **C22-26** | **>= 105** | 0.39 (0.15-1.04) | 0.06 | 2.27 (0.78-6.64) | 0.13 | 6.13 (1.62-23.2) | 0.01 |
|  | **90 - <105** | 0.69 (0.42-1.14) | 0.15 | 1.49 (0.91-2.45) | 0.12 | 2.10 (1.10-4.03) | 0.03 |
|  | **75 - <90** | Ref | NA | Ref | NA | Ref | NA |
|  | **60 - <75** | 0.89 (0.55-1.45) | 0.64 | 0.92 (0.57-1.49) | 0.73 | 0.98 (0.50-1.92) | 0.95 |
|  | **45 - <60** | 0.89 (0.49-1.62) | 0.69 | 1.06 (0.57-1.98) | 0.85 | 1.11 (0.48-2.57) | 0.81 |
|  | **<45** | 0.56 (0.31-1.04) | 0.06 | 0.78 (0.40-1.54) | 0.47 | 1.26 (0.53-2.98) | 0.60 |
| **Digestive tract** | **P for trend** |  | Linear 0.84<br>Non-linear 0.85 |  | Linear 0.04<br>Non-linear 0.01 |  | Linear 0.27<br>Non-linear 0.12 |
| **C15-21** | **>= 105** | 1.41 (0.79-2.50) | 0.24 | 2.04 (1.29-3.21) | 0.002 | 1.35 (0.67-2.71) | 0.40 |
|  | **90 - <105** | 1.04 (0.83-1.29) | 0.76 | 1.18 (0.99-1.40) | 0.06 | 1.11 (0.85-1.44) | 0.45 |
|  | **75 - <90** | Ref | NA | Ref | NA | Ref | NA |
|  | **60 - <75** | 1.25 (1.04-1.51) | 0.02 | 1.10 (0.94-1.28) | 0.24 | 0.88 (0.69-1.12) | 0.30 |
|  | **45 - <60** | 1.30 (1.03-1.65) | 0.03 | 1.09 (0.90-1.32) | 0.39 | 0.84 (0.63-1.13) | 0.25 |
|  | **<45** | 0.96 (0.74-1.24) | 0.75 | 0.98 (0.79-1.21) | 0.83 | 1.03 (0.75-1.41) | 0.87 |
| **Head and neck** | **P for trend** |  | Linear 0.73<br>Non-linear 0.04 |  | Linear 0.03<br>Non-linear 0.51 |  | Linear 0.05<br>Non-linear 0.24 |
| **C00-14** | **>= 105** | 2.09 (0.81-5.44) | 0.13 | 1.96 (1.00-3.84) | 0.05 | 1.08 (0.38-3.04) | 0.89 |
| **C30-32** | **90 - <105** | 2.08 (1.20-3.60) | 0.01 | 1.24 (0.89-1.72) | 0.21 | 0.59 (0.33-1.05) | 0.07 |
|  | **75 - <90** | Ref | NA | Ref | NA | Ref | NA |
|  | **60 - <75** | 2.08 (1.10-3.95) | 0.02 | 1.14 (0.79-1.66) | 0.48 | 0.54 (0.26-1.11) | 0.09 |
|  | **45 - <60** | 0.86 (0.41-1.80) | 0.69 | 1.00 (0.57-1.73) | 0.99 | 1.07 (0.45-2.58) | 0.87 |
|  | **<45** | 2.20 (0.74-6.54) | 0.16 | 0.57 (0.31-1.07) | 0.08 | 0.25 (0.08-0.83) | 0.02 |
| **Lung** | **P for trend** |  | Linear 0.04<br>Non-linear 0.04 |  | Linear 0.90<br>Non-linear 0.92 |  | Linear 0.26<br>Non-linear 0.26 |
| **C33-34** | **>= 105** | 1.93 (1.03-3.61) | 0.04 | 1.17 (0.70-1.96) | 0.54 | 0.73 (0.33-1.59) | 0.43 |
|  | **90 - <105** | 1.17 (0.93-1.46) | 0.18 | 1.09 (0.87-1.36) | 0.47 | 1.01 (0.75-1.36) | 0.95 |
|  | **75 - <90** | Ref | NA | Ref | NA | Ref | NA |
|  | **60 - <75** | 0.93 (0.76-1.14) | 0.50 | 1.23 (1.00-1.52) | 0.05 | 1.29 (0.96-1.73) | 0.09 |
|  | **45 - <60** | 0.99 (0.78-1.27) | 0.95 | 1.14 (0.89-1.47) | 0.29 | 1.09 (0.78-1.53) | 0.60 |
|  | **<45** | 0.90 (0.69-1.19) | 0.47 | 1.02 (0.78-1.34) | 0.87 | 1.06 (0.73-1.54) | 0.75 |
| **Melanoma** | **P for trend** |  | Linear 0.48<br>Non-linear 0.98 |  | Linear 0.85<br>Non-linear 0.80 |  | Linear 0.42<br>Non-linear 0.96 |
| **C43** | **>= 105** | 1.99 (0.55-7.24) | 0.30 | 0.71 (0.18-2.86) | 0.63 | 0.40 (0.08-2.05) | 0.27 |
|  | **90 - <105** | 1.57 (0.71-3.44) | 0.26 | 0.56 (0.28-1.13) | 0.11 | 0.35 (0.13-0.94) | 0.04 |
|  | **75 - <90** | Ref | NA | Ref | NA | Ref | NA |
|  | **60 - <75** | 1.69 (0.84-3.43) | 0.14 | 0.63 (0.35-1.13) | 0.12 | 0.35 (0.14-0.87) | 0.02 |
|  | **45 - <60** | 1.40 (0.54-3.62) | 0.48 | 0.84 (0.43-1.66) | 0.62 | 0.61 (0.20-1.83) | 0.38 |
|  | **<45** | 0.64 (0.17-2.36) | 0.50 | 1.01 (0.45-2.26) | 0.98 | 1.66 (0.38-7.27) | 0.50 |
| **Other** | **P for trend** |  | Linear 0.61<br>Non-linear 0.46 |  | Linear 0.40<br>Non-linear 0.12 |  | Linear 0.58<br>Non-linear 0.28 |
| **C37-38** | **>= 105** | 8.00 (0.80-80.0) | 0.08 | 3.04 (0.26-35.5) | 0.38 | 1.24 (0.06-25.5) | 0.89 |
| **C45-49** | **90 - <105** | 0.91 (0.27-3.04) | 0.87 | 1.19 (0.54-2.60) | 0.66 | 2.12 (0.58-7.72) | 0.25 |
| **C69-72** | **75 - <90** | Ref | NA | Ref | NA | Ref | NA |
|  | **60 - <75** | 0.71 (0.22-2.27) | 0.56 | 0.87 (0.43-1.77) | 0.70 | 1.36 (0.37-4.97) | 0.65 |
|  | **45 - <60** | 2.91 (0.52-16.3) | 0.22 | 0.48 (0.19-1.21) | 0.12 | 0.25 (0.04-1.44) | 0.12 |
|  | **<45** | 0.98 (0.17-5.76) | 0.98 | 1.28 (0.34-4.81) | 0.72 | 1.25 (0.16-9.90) | 0.83 |
| **Renal tract** | **P for trend** |  | Linear 0.17<br>Non-linear 0.44 |  | Linear 0.03<br>Non-linear 0.33 |  | Linear 0.85<br>Non-linear 0.75 |
| **C64-67** | **>= 105** | 1.05 (0.33-3.35) | 0.94 | 0.81 (0.43-1.53) | 0.52 | 0.82 (0.24-2.86) | 0.75 |
|  | **90 - <105** | 1.34 (0.81-2.20) | 0.26 | 0.78 (0.55-1.12) | 0.18 | 0.61 (0.34-1.09) | 0.10 |
|  | **75 - <90** | Ref | NA | Ref | NA | Ref | NA |
|  | **60 - <75** | 1.56 (0.99-2.46) | 0.06 | 1.04 (0.76-1.43) | 0.81 | 0.66 (0.38-1.15) | 0.15 |
|  | **45 - <60** | 1.94 (1.24-3.03) | 0.004 | 1.26 (0.92-1.74) | 0.15 | 0.64 (0.38-1.09) | 0.10 |
|  | **<45** | 1.32 (0.81-2.13) | 0.26 | 1.39 (0.99-1.95) | 0.05 | 1.04 (0.60-1.80) | 0.89 |
| **Breast** | **P for trend** |  | Linear 0.01<br>Non-linear 0.06 |  | NA |  | NA |
| **C50** | **>= 105** | 1.65 (1.12-2.44) | 0.01 | NA | NA | NA | NA |
|  | **90 - <105** | 0.80 (0.65-0.99) | 0.04 | NA | NA | NA | NA |
|  | **75 - <90** | Ref | NA | NA | NA | NA | NA |
|  | **60 - <75** | 1.08 (0.89-1.32) | 0.43 | NA | NA | NA | NA |
|  | **45 - <60** | 1.38 (1.07-1.79) | 0.01 | NA | NA | NA | NA |
|  | **<45** | 1.95 (1.44-2.63) | 0 | NA | NA | NA | NA |
| **Female genital tract** | **P for trend** |  | Linear 0.17<br>Non-linear 0.60 |  | NA |  | NA |
| **C51-58** | **>= 105** | 1.00 (0.62-1.61) | 1.00 | NA | NA | NA | NA |
|  | **90 - <105** | 0.93 (0.73-1.20) | 0.59 | NA | NA | NA | NA |
|  | **75 - <90** | Ref | NA | NA | NA | NA | NA |
|  | **60 - <75** | 0.84 (0.67-1.06) | 0.13 | NA | NA | NA | NA |
|  | **45 - <60** | 1.16 (0.88-1.53) | 0.29 | NA | NA | NA | NA |
|  | **<45** | 1.41 (1.01-1.99) | 0.05 | NA | NA | NA | NA |
| **Prostate** | **P for trend** |  | NA |  | Linear 0.96<br>Non-linear <0.001 |  | NA |
| **C61** | **>= 105** | NA | NA | 2.02 (1.16-3.51) | 0.01 | NA | NA |
|  | **90 - <105** | NA | NA | 1.34 (1.16-1.56) | <0.001 | NA | NA |
|  | **75 - <90** | NA | NA | Ref | NA | NA | NA |
|  | **60 - <75** | NA | NA | 1.03 (0.90-1.17) | 0.66 | NA | NA |
|  | **45 - <60** | NA | NA | 1.13 (0.95-1.34) | 0.17 | NA | NA |
|  | **<45** | NA | NA | 1.48 (1.20-1.82) | <0.001 | NA | NA |

Logistic regression models adjusted for age, deprivation status, smoking status, comorbidity count and cancer site. “Female” and “Male” models are stratified by sex. “Male versus female” model includes and interaction term between eGFR category and sex. Linear: P for linear trend. Non-linear: P for cubic trend. eGFR: estimated glomerular filtration rate based on CKD-EPI 2009 equation and using serum creatinine. OR: odds ratio. CI: confidence interval.

**Table S4.** Hazards of all-cause mortality across all sites by eGFR category.

|  | Female |  | Male |  | Male versus female |  |
| --- | --- | --- | --- | --- | --- | --- |
| eGFR category<br>(mL/min/1.73m<sup>2</sup>) | HR (95% CI) | P value | HR (95% CI) | P value | HR (95% CI) | P value |
| P for trend |  | Linear 0.16<br>Non-linear <0.001 |  | Linear <0.001<br>Non-linear <0.001 |  | Linear 0.27<br>Non-linear <0.001 |
| >120 | 1.52 (0.93-2.50) | 0.10 | 2.92 (2.21-3.85) | <0.001 | 1.80 (1.02-3.16) | 0.04 |
| 105 - <120 | 1.63 (1.42-1.85) | <0.001 | 1.81 (1.63-2.02) | <0.001 | 1.06 (0.91-1.24) | 0.46 |
| 90 - <105 | 1.17 (1.10-1.24) | <0.001 | 1.14 (1.08-1.20) | <0.001 | 0.95 (0.89-1.02) | 0.17 |
| 75 - <90 | Ref | NA | Ref | NA | Ref | NA |
| 60 - <75 | 1.09 (1.03-1.14) | <0.001 | 1.03 (0.99-1.08) | 0.13 | 0.96 (0.90-1.02) | 0.17 |
| 45 - <60 | 1.19 (1.13-1.26) | <0.001 | 1.12 (1.07-1.18) | <0.001 | 0.96 (0.89-1.03) | 0.24 |
| 30 - <45 | 1.30 (1.21-1.38) | <0.001 | 1.17 (1.11-1.25) | <0.001 | 0.93 (0.85-1.01) | 0.07 |
| <30 | 1.71 (1.56-1.88) | <0.001 | 1.48 (1.36-1.60) | <0.001 | 0.88 (0.78-0.99) | 0.04 |

Cox proportional hazards models adjusted for age, deprivation status, smoking status, comorbidity count, cancer site and presenting cancer stage. eGFR: estimated glomerular filtration rate based on CKD-EPI 2009 equation and using serum creatinine. “Female” and “Male” models are stratified by sex. “Male versus female” model includes an interaction term between eGFR category and sex. Linear: P for linear trend. Non-linear: P for cubic trend. HR: hazards ratio. CI: confidence interval.

**Table S5.**
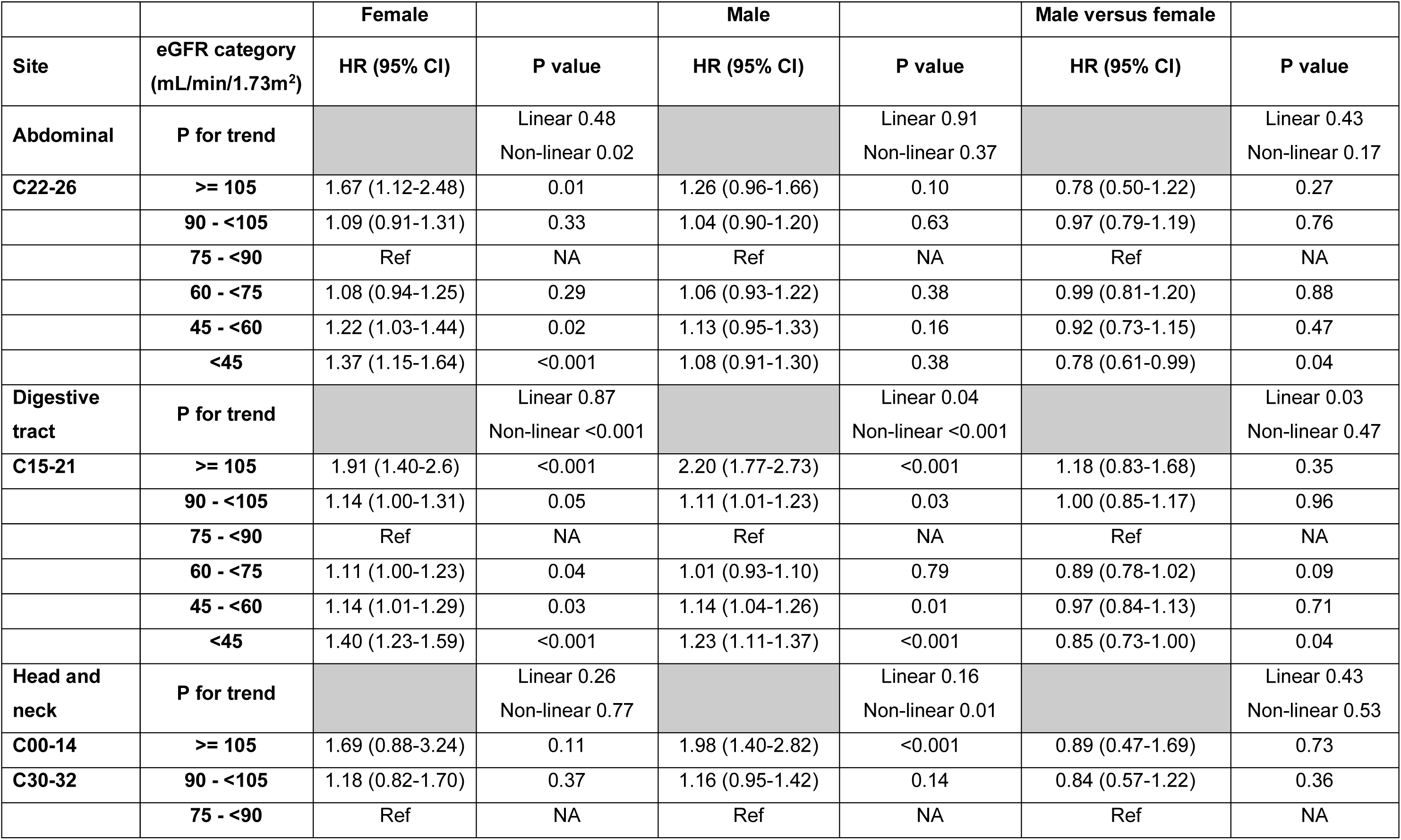
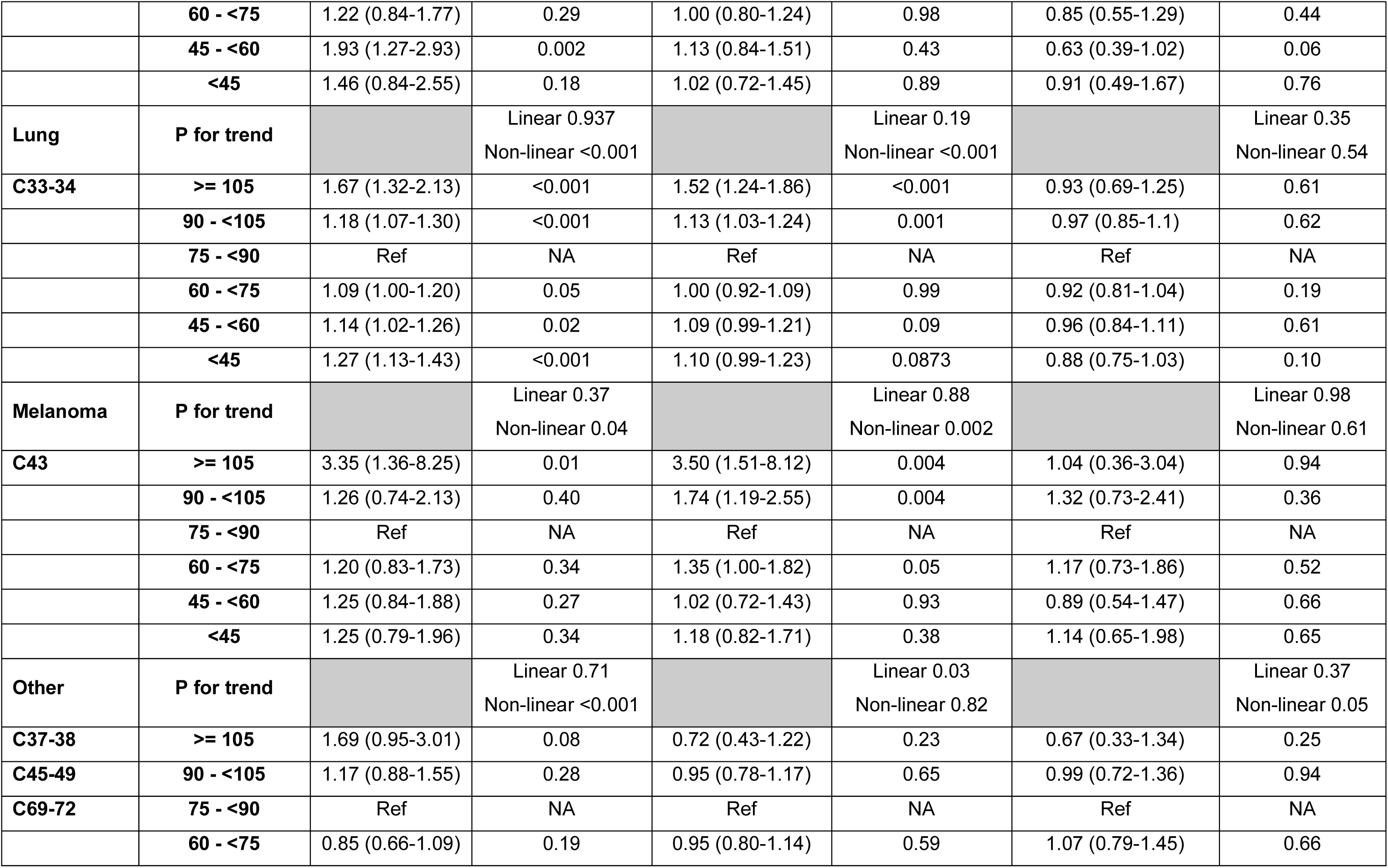
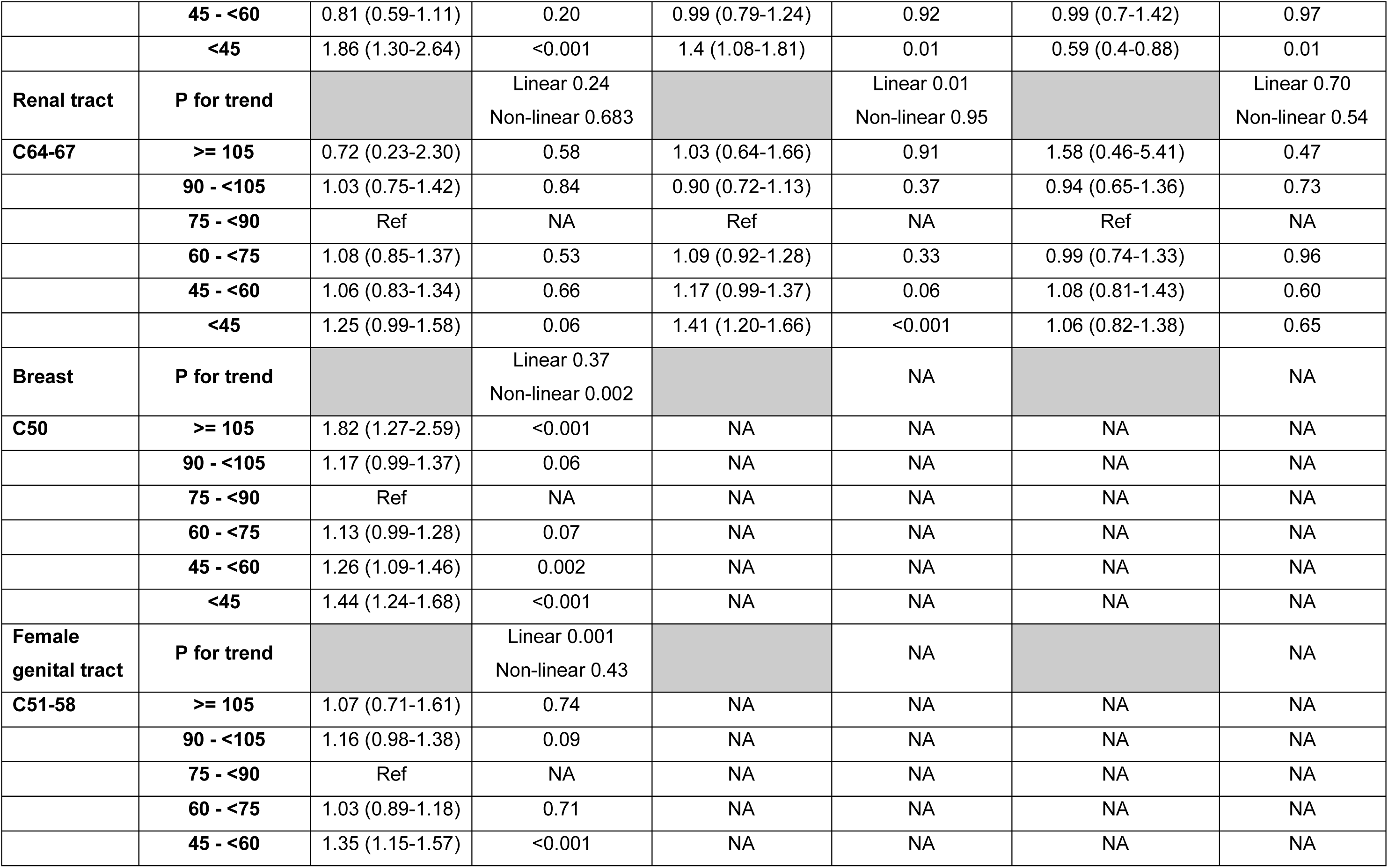
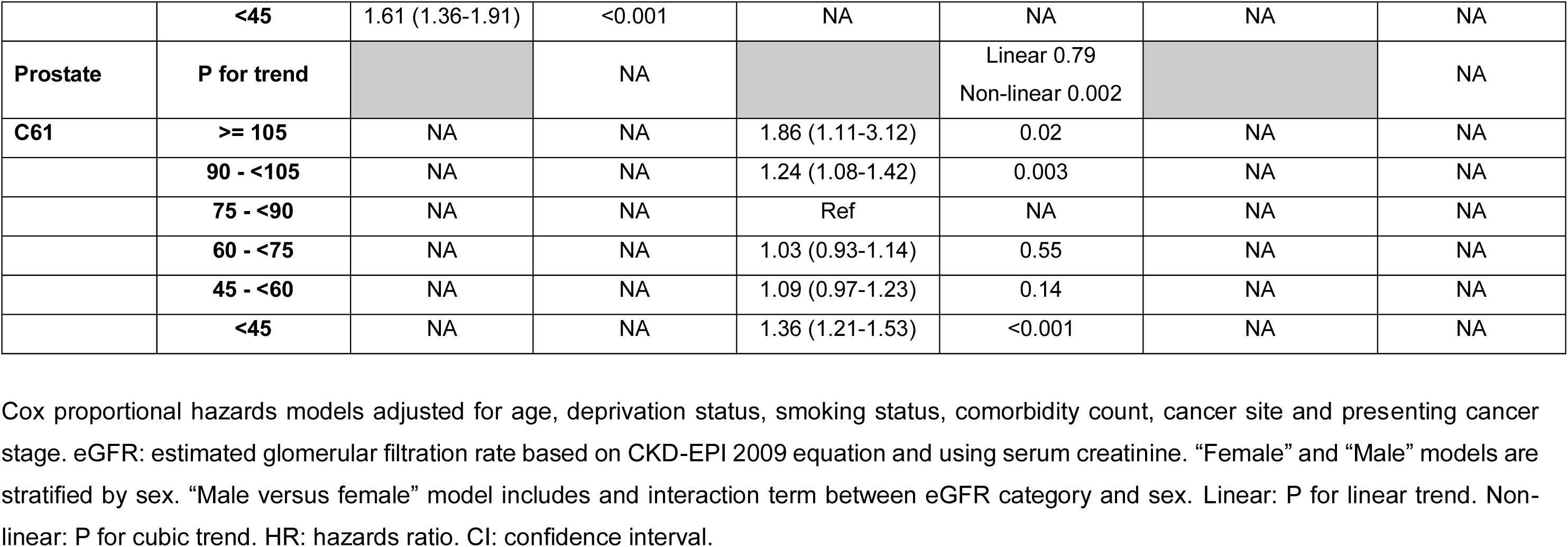
Hazards of all-cause mortality by cancer site and eGFR category.

|  |  | Female |  | Male |  | Male versus female |  |
| --- | --- | --- | --- | --- | --- | --- | --- |
| Site | eGFR category<br>(mL/min/1.73m<sup>2</sup>) | HR (95% CI) | P value | HR (95% CI) | P value | HR (95% CI) | P value |
| **Abdominal** | **P for trend** |  | Linear 0.48<br>Non-linear 0.02 |  | Linear 0.91<br>Non-linear 0.37 |  | Linear 0.43<br>Non-linear 0.17 |
| **C22-26** | **>= 105** | 1.67 (1.12-2.48) | 0.01 | 1.26 (0.96-1.66) | 0.10 | 0.78 (0.50-1.22) | 0.27 |
|  | **90 - <105** | 1.09 (0.91-1.31) | 0.33 | 1.04 (0.90-1.20) | 0.63 | 0.97 (0.79-1.19) | 0.76 |
|  | **75 - <90** | Ref | NA | Ref | NA | Ref | NA |
|  | **60 - <75** | 1.08 (0.94-1.25) | 0.29 | 1.06 (0.93-1.22) | 0.38 | 0.99 (0.81-1.20) | 0.88 |
|  | **45 - <60** | 1.22 (1.03-1.44) | 0.02 | 1.13 (0.95-1.33) | 0.16 | 0.92 (0.73-1.15) | 0.47 |
|  | **<45** | 1.37 (1.15-1.64) | <0.001 | 1.08 (0.91-1.30) | 0.38 | 0.78 (0.61-0.99) | 0.04 |
| **Digestive tract** | **P for trend** |  | Linear 0.87<br>Non-linear <0.001 |  | Linear 0.04<br>Non-linear <0.001 |  | Linear 0.03<br>Non-linear 0.47 |
| **C15-21** | **>= 105** | 1.91 (1.40-2.6) | <0.001 | 2.20 (1.77-2.73) | <0.001 | 1.18 (0.83-1.68) | 0.35 |
|  | **90 - <105** | 1.14 (1.00-1.31) | 0.05 | 1.11 (1.01-1.23) | 0.03 | 1.00 (0.85-1.17) | 0.96 |
|  | **75 - <90** | Ref | NA | Ref | NA | Ref | NA |
|  | **60 - <75** | 1.11 (1.00-1.23) | 0.04 | 1.01 (0.93-1.10) | 0.79 | 0.89 (0.78-1.02) | 0.09 |
|  | **45 - <60** | 1.14 (1.01-1.29) | 0.03 | 1.14 (1.04-1.26) | 0.01 | 0.97 (0.84-1.13) | 0.71 |
|  | **<45** | 1.40 (1.23-1.59) | <0.001 | 1.23 (1.11-1.37) | <0.001 | 0.85 (0.73-1.00) | 0.04 |
| **Head and neck** | **P for trend** |  | Linear 0.26<br>Non-linear 0.77 |  | Linear 0.16<br>Non-linear 0.01 |  | Linear 0.43<br>Non-linear 0.53 |
| **C00-14** | **>= 105** | 1.69 (0.88-3.24) | 0.11 | 1.98 (1.40-2.82) | <0.001 | 0.89 (0.47-1.69) | 0.73 |
| **C30-32** | **90 - <105** | 1.18 (0.82-1.70) | 0.37 | 1.16 (0.95-1.42) | 0.14 | 0.84 (0.57-1.22) | 0.36 |
|  | **75 - <90** | Ref | NA | Ref | NA | Ref | NA |
|  | **60 - <75** | 1.22 (0.84-1.77) | 0.29 | 1.00 (0.80-1.24) | 0.98 | 0.85 (0.55-1.29) | 0.44 |
|  | **45 - <60** | 1.93 (1.27-2.93) | 0.002 | 1.13 (0.84-1.51) | 0.43 | 0.63 (0.39-1.02) | 0.06 |
|  | **<45** | 1.46 (0.84-2.55) | 0.18 | 1.02 (0.72-1.45) | 0.89 | 0.91 (0.49-1.67) | 0.76 |
| **Lung** | **P for trend** |  | Linear 0.937<br>Non-linear <0.001 |  | Linear 0.19<br>Non-linear <0.001 |  | Linear 0.35<br>Non-linear 0.54 |
| **C33-34** | **>= 105** | 1.67 (1.32-2.13) | <0.001 | 1.52 (1.24-1.86) | <0.001 | 0.93 (0.69-1.25) | 0.61 |
|  | **90 - <105** | 1.18 (1.07-1.30) | <0.001 | 1.13 (1.03-1.24) | 0.001 | 0.97 (0.85-1.1) | 0.62 |
|  | **75 - <90** | Ref | NA | Ref | NA | Ref | NA |
|  | **60 - <75** | 1.09 (1.00-1.20) | 0.05 | 1.00 (0.92-1.09) | 0.99 | 0.92 (0.81-1.04) | 0.19 |
|  | **45 - <60** | 1.14 (1.02-1.26) | 0.02 | 1.09 (0.99-1.21) | 0.09 | 0.96 (0.84-1.11) | 0.61 |
|  | **<45** | 1.27 (1.13-1.43) | <0.001 | 1.10 (0.99-1.23) | 0.0873 | 0.88 (0.75-1.03) | 0.10 |
| **Melanoma** | **P for trend** |  | Linear 0.37<br>Non-linear 0.04 |  | Linear 0.88<br>Non-linear 0.002 |  | Linear 0.98<br>Non-linear 0.61 |
| **C43** | **>= 105** | 3.35 (1.36-8.25) | 0.01 | 3.50 (1.51-8.12) | 0.004 | 1.04 (0.36-3.04) | 0.94 |
|  | **90 - <105** | 1.26 (0.74-2.13) | 0.40 | 1.74 (1.19-2.55) | 0.004 | 1.32 (0.73-2.41) | 0.36 |
|  | **75 - <90** | Ref | NA | Ref | NA | Ref | NA |
|  | **60 - <75** | 1.20 (0.83-1.73) | 0.34 | 1.35 (1.00-1.82) | 0.05 | 1.17 (0.73-1.86) | 0.52 |
|  | **45 - <60** | 1.25 (0.84-1.88) | 0.27 | 1.02 (0.72-1.43) | 0.93 | 0.89 (0.54-1.47) | 0.66 |
|  | **<45** | 1.25 (0.79-1.96) | 0.34 | 1.18 (0.82-1.71) | 0.38 | 1.14 (0.65-1.98) | 0.65 |
| **Other** | **P for trend** |  | Linear 0.71<br>Non-linear <0.001 |  | Linear 0.03<br>Non-linear 0.82 |  | Linear 0.37<br>Non-linear 0.05 |
| **C37-38** | **>= 105** | 1.69 (0.95-3.01) | 0.08 | 0.72 (0.43-1.22) | 0.23 | 0.67 (0.33-1.34) | 0.25 |
| **C45-49** | **90 - <105** | 1.17 (0.88-1.55) | 0.28 | 0.95 (0.78-1.17) | 0.65 | 0.99 (0.72-1.36) | 0.94 |
| **C69-72** | **75 - <90** | Ref | NA | Ref | NA | Ref | NA |
|  | **60 - <75** | 0.85 (0.66-1.09) | 0.19 | 0.95 (0.80-1.14) | 0.59 | 1.07 (0.79-1.45) | 0.66 |
|  | **45 - <60** | 0.81 (0.59-1.11) | 0.20 | 0.99 (0.79-1.24) | 0.92 | 0.99 (0.7-1.42) | 0.97 |
|  | **<45** | 1.86 (1.30-2.64) | <0.001 | 1.4 (1.08-1.81) | 0.01 | 0.59 (0.4-0.88) | 0.01 |
| **Renal tract** | **P for trend** |  | Linear 0.24<br>Non-linear 0.683 |  | Linear 0.01<br>Non-linear 0.95 |  | Linear 0.70<br>Non-linear 0.54 |
| **C64-67** | **>= 105** | 0.72 (0.23-2.30) | 0.58 | 1.03 (0.64-1.66) | 0.91 | 1.58 (0.46-5.41) | 0.47 |
|  | **90 - <105** | 1.03 (0.75-1.42) | 0.84 | 0.90 (0.72-1.13) | 0.37 | 0.94 (0.65-1.36) | 0.73 |
|  | **75 - <90** | Ref | NA | Ref | NA | Ref | NA |
|  | **60 - <75** | 1.08 (0.85-1.37) | 0.53 | 1.09 (0.92-1.28) | 0.33 | 0.99 (0.74-1.33) | 0.96 |
|  | **45 - <60** | 1.06 (0.83-1.34) | 0.66 | 1.17 (0.99-1.37) | 0.06 | 1.08 (0.81-1.43) | 0.60 |
|  | **<45** | 1.25 (0.99-1.58) | 0.06 | 1.41 (1.20-1.66) | <0.001 | 1.06 (0.82-1.38) | 0.65 |
| **Breast** | **P for trend** |  | Linear 0.37<br>Non-linear 0.002 |  | NA |  | NA |
| **C50** | **>= 105** | 1.82 (1.27-2.59) | <0.001 | NA | NA | NA | NA |
|  | **90 - <105** | 1.17 (0.99-1.37) | 0.06 | NA | NA | NA | NA |
|  | **75 - <90** | Ref | NA | NA | NA | NA | NA |
|  | **60 - <75** | 1.13 (0.99-1.28) | 0.07 | NA | NA | NA | NA |
|  | **45 - <60** | 1.26 (1.09-1.46) | 0.002 | NA | NA | NA | NA |
|  | **<45** | 1.44 (1.24-1.68) | <0.001 | NA | NA | NA | NA |
| **Female genital tract** | **P for trend** |  | Linear 0.001<br>Non-linear 0.43 |  | NA |  | NA |
| **C51-58** | **>= 105** | 1.07 (0.71-1.61) | 0.74 | NA | NA | NA | NA |
|  | **90 - <105** | 1.16 (0.98-1.38) | 0.09 | NA | NA | NA | NA |
|  | **75 - <90** | Ref | NA | NA | NA | NA | NA |
|  | **60 - <75** | 1.03 (0.89-1.18) | 0.71 | NA | NA | NA | NA |
|  | **45 - <60** | 1.35 (1.15-1.57) | <0.001 | NA | NA | NA | NA |
|  | **<45** | 1.61 (1.36-1.91) | <0.001 | NA | NA | NA | NA |
| **Prostate** | **P for trend** |  | NA |  | Linear 0.79<br>Non-linear 0.002 |  | NA |
| **C61** | **>= 105** | NA | NA | 1.86 (1.11-3.12) | 0.02 | NA | NA |
|  | **90 - <105** | NA | NA | 1.24 (1.08-1.42) | 0.003 | NA | NA |
|  | **75 - <90** | NA | NA | Ref | NA | NA | NA |
|  | **60 - <75** | NA | NA | 1.03 (0.93-1.14) | 0.55 | NA | NA |
|  | **45 - <60** | NA | NA | 1.09 (0.97-1.23) | 0.14 | NA | NA |
|  | **<45** | NA | NA | 1.36 (1.21-1.53) | <0.001 | NA | NA |

Cox proportional hazards models adjusted for age, deprivation status, smoking status, comorbidity count, cancer site and presenting cancer stage. eGFR: estimated glomerular filtration rate based on CKD-EPI 2009 equation and using serum creatinine. “Female” and “Male” models are stratified by sex. “Male versus female” model includes an interaction term between eGFR category and sex. Linear: P for linear trend. Non-linear: P for cubic trend. HR: hazards ratio. CI: confidence interval.

